# Molecular Methods to Detect *Vibrio cholerae* and Associated Bacteriophages among Diarrheal Patients in Bangladesh

**DOI:** 10.64898/2026.05.08.26352758

**Authors:** I. Sriguha, M. Mu, Abu Sayeed, E. T. Cato, A. Creasy-Marrazzo, K. Islam, I.UL. Khabir, T. R. Bhuiyan, Y. Begum, Taufiqul Islam, Zahid Hasan Khan, E. Freeman, A. Vustepalli, L. Brinkley, D. G. Brown, D.J. Pouchnik, K. Mi, Z. Lin, J.A. Grembi, D.T. Leung, F. Qadri, A. I. Khan, E. J. Nelson

## Abstract

Molecular diagnostics to detect *Vibrio cholerae* (*Vc*) may be negatively impacted by pathogen-specific lytic bacteriophage (phage) predation. To address this problem, phage detection as a proxy for pathogen detection has been proposed. However, efforts to modernize cholera diagnostics with molecular tools require addressing knowledge gaps on best practices to detect *Vc* and associated bacteriophages. We conducted polymerase chain reaction (PCR), quantitative PCR (qPCR), and nano-liter (nl) qPCR targeting *Vc* and known phages (ICP1/2/3) on stool samples collected from patients admitted at hospitals across Bangladesh. Of 4,975 patients enrolled, 2,574 diarrheal samples were collected and over 65,000 reactions were conducted, including replicates. We analyzed the results for target-specific assay alignment and then used machine learning to determine the effect of phage predation on *Vc*-assay alignment. Standard curve analyses were used to set qPCR-positivity thresholds at 7.3×10^5^ CFU/mL for *Vc* and 1.7×10^3^, 9.3×10^3^, and 3.0×10^5^ PFU/mL for ICP1, ICP2, and ICP3, respectively. Among 2,462 samples assayed by qPCR, target detection was 25.3% (623), 7.8% (193), 0.5% (13), and 5.8% (144) for *Vc*, ICP1, ICP2, and ICP3, respectively. There was strong alignment between assays for *Vc* detection (κ=0.785) and moderate alignment for phage detection (κ=0.609, 0.593, and 0.533 for ICP1/2/3, respectively). Phages were ranked as the first (ICP1) and third (ICP3) effectors of *Vc* diagnostic alignment. These findings provide insights on how to prioritize molecular methods in the cholera field as well as related less tractable diseases facing similar diagnostic challenges.

**IMPORTANCE:** This paper presents a comprehensive comparison of molecular methods to detect *Vibrio cholerae* (*Vc*) and associated bacteriophage (phage) which can be used as a proxy for pathogen detection. This initiative is an important step towards modernizing cholera diagnostics with molecular tools. In this study, we found that quantitative polymerase chain reaction (qPCR) represents a reasonable approach to detect *Vc* and associated phages balancing assay performance, cost, and accessibility. A key additional finding was that phage predation was found to be a leading factor that impacts the alignment of molecular methods to detect *Vc*. While we recommend qPCR be added to the cholera diagnostic toolkit, the effects of phage predation need to be accounted for in the development and evaluation of cholera diagnostics. These findings have applicability to less tractable disease where diagnostics share similar vulnerabilities.

## INTRODUCTION

Cholera is a diarrheal disease caused by the Gram-negative water-borne pathogen *Vibrio cholerae* (*Vc*). Globally, there are 1.3 to 4 million reported cases annually resulting in 21,000 to 143,000 deaths (1). Methods to detect *Vc* and related enteric pathogens include culture, polymerase chain reaction (PCR) and quantitative PCR (qPCR). Nano-liter qPCR (nl-qPCR) is a scalable alternative to conventional qPCR that utilizes a high-throughput platform of nanofluidic volumes (2) and can reduce cost of *Vc* detection six-fold (3). PCR-based molecular diagnostics generally offer faster, more sensitive and more specific results than culture (4, 5).

Molecular diagnostics have limitations and vulnerabilities. These vulnerabilities include the effects of antimicrobial agents simply reducing the target number below the limit of detection. Furthermore, virulent bacteriophages (phage) encode nucleases that can rapidly degrade target *Vc* nucleic acid (6, 7). This degradation can compromise PCR-based assays (8) and poses a risk of false negative results (9, 10). Accurate, low-cost and expedient diagnostics are critical to cholera outbreak response: they decrease time-to-treatment while ensuring antibiotic stewardship; they are important to effective cholera outbreak response (11). However, deciding which molecular diagnostic approach to deploy is complex, especially in resource-limited settings with barriers to accessing reagents, machinery and expertise (12, 13).

To develop a rationale for selecting a molecular diagnostic panel for cholera, we sought to address two knowledge gaps: (i) the degree of alignment between PCR-based molecular assays for the detection of *Vc* and associated virulent lytic phage (ICP1, ICP2, and ICP3); and (ii) the factors that impact the alignment of molecular assays for *Vc*. In this study, we conducted PCR, qPCR, and nl-qPCR targeting *Vc* and associated phages on samples collected from patients admitted at hospitals across Bangladesh. We analyzed the dataset for alignment by assay type, assessed the relative impact of phage predation as an effector of *Vc* diagnostic alignment, and performed a comparative cost analysis to assist decisions on which diagnostic approach to take in different settings.

## METHODS

### Ethics Statement

Research ethics boards at the Institute of Epidemiology, Disease Control and Research (IEDCR), International Centre for Diarrhoeal Disease Research, Bangladesh (icddr,b) and the University of Florida (UF) approved the mHealth Diarrhea Management (mHDM) cluster randomized controlled trial conducted in 2018-2019 (IEDCR IRB/2017/10; icddr,b ERC/RRC PR-17036; University of Florida IRB 201601762) (14). Research ethics boards at the icddr,b approved the National Cholera Surveillance (icddr,b ERC/RRC PR-15127) study (15). Informed consent was obtained from adult participants or the parents/legal guardians of children for the National Cholera Surveillance (NCS) study. Participants between 11 years and under 18 years provided written assent (14). Parents/legal guardians of children under the age of 11 provided written consent. This study represents a secondary analysis on data and samples collected in a previously published parent study (14). The study objectives are within the scope and intent of the original study and associated consent/assent documents.

### Clinical Procedures

A cluster randomized controlled trial was conducted in the parent study, enrolling patients with acute diarrheal disease across ten district hospitals in Bangladesh (Fig S1) (14). As etiology of diarrheal disease differs between children and adults (16, 17), stool sample collection targeted two groups: children under 5 years of age and individuals older than 5 years. Stool samples were collected daily from two patients within each age group. If collecting two samples from an age group was not possible, over-enrollment was done for the other group to ensure that a total of four samples were collected per day. Field samples were aliquoted in Cary-Blair transport media to culture using standard methods icddr,b (14, 18). and in RNAlater™ (Invitrogen) for total nucleic acid (tNA) extraction using standard methods at UF (18).

Patients’ demographic data including sex, age, and sample site were recorded (14). Clinical symptoms of vomiting, duration of diarrhea, dehydration status, and self-reported antibiotic use were reported and used for analysis in this study.

### Laboratory procedures

#### PCR assay

PCR was performed on the tNA targeting the following: an outer membrane protein (*ompW*) for *Vc* (19), a uniquely encoded DNA polymerase I gene product (GP58) for ICP1 (20), a hypothetical protein (GP4) and a putative tail fiber protein (GP24) for ICP2 (21, 22), and a hypothetical protein (GP5) for ICP3 (20) (Table S1). *Vc* (*tcpA*) positive and negative DNA template controls were run alongside samples (Fig S2). A 20 µL PCR master mix was prepared with 10 µL of OneTaq 2x Master Mix with Taq and Deep Vent polymerases from New England Biolabs or 10 µL of MyTaq HS Red Mix with a Taq polymerase from Meridian Biosciences, 1 µL of the 10 µM forward and reverse primer mix, 9 µL of nuclease-free water (NFW), and 1 µL of the tNA sample in each well. Confirmatory PCR for phage were prepared with 50 µL of master mix consisting of 25 µL of Q5 High-Fidelity 2X Master Mix (NEB), 2 µL of 10 µM forward and reverse primer mix, 23 µL of NFW, and 1 µL of tNA sample. Targets were amplified on a Bio-Rad T100 Thermal Cycler. For *Vc*, samples were denatured at 95 °C for 2 minutes followed by 35 cycles of denaturation at 95 °C for 45 seconds, annealing at 51 °C for 30 seconds, and extension at 72 °C for 1 minute. A final 10-minute extension step was completed at 72 °C. For ICP1, samples were denatured at 94 °C for 30 seconds followed by 35 cycles of denaturation at 94 °C for 30 seconds, annealing at 51 °C for 30 seconds, and extension at 68 °C for one minute. A final extension for 5 minutes was completed at 68 °C. ICP2 and ICP3 PCR parameters were similar with annealing temperatures of 56 °C and 54 °C, respectively. Resulting amplicons were run on 1.5% agarose gels (Lonza SeaKem LE Agarose) at 120-140 volts and visualized with Invitrogen SYBR Safe™ DNA Gel Stain (Fig S2). Samples positive for phage were confirmed with a biological replicate. A subset of samples targeting *Vc* (phage positive samples and 10% of culture and phage negative samples) were assayed in two biological replicates. Conflicting results were resolved by a third replicate.

#### qPCR assay

Quantitative PCR-specific primer sets were used to target the *tcpA* (toxin co-regulated pilus), GP58, and GP19 genes of *Vc*, ICP1, and ICP3, respectively (Table S1) (18, 23). A generalized primer set for PCR and qPCR was used to target the gene GP24 of ICP2 (21). A primer set to *16S* rDNA gene (24) was used as eubacterial control to assess run failures. Reagents were mixed at the same ratios as PCR using NEB Luna Universal qPCR Master Mix. One microliter of tNA sample was loaded in each well of a 96-well plate along with pre-prepared master mix. *Vc* (*tcpA*) positive and negative DNA template controls were run alongside samples (Fig S3). Targets were amplified on a Bio Rad CFX96 Touch Real-Time PCR Detection System. For the *tcpA* primers, samples were denatured at 95 °C for one minute followed by 35 cycles of denaturation at 95 °C for 15 seconds, annealing at 60 °C for 30 seconds, and extension at 72 °C for one minute. For ICP1, ICP3, and *16S* rDNA primers, samples were denatured at 98 °C for one minute followed by 35 cycles of denaturation at 98 °C for 15 seconds and annealing at 60 °C for 30 seconds. For the ICP2 primers, samples were denatured at 98 °C for one minute followed by 35 cycles of denaturation at 98 °C for 15 seconds and annealing at 63 °C for 30 seconds.

#### nl-qPCR assay

Nanoliter qPCR (nl-qPCR) was conducted for the same targets (*tcpA*, GP58, GP24, and GP19 genes for *Vc*, ICP1, ICP2, and ICP3, respectively) as qPCR (Table S1). A gBlock-based standard curve specific to nl-qPCR was included as an internal plate control for the same target genes; the gBlocks were commercially purchased from Integrated DNA Technologies (IDT). A starting concentration of 5 x 10^7^ copies of DNA/μL was diluted in a series of seven 10-fold dilutions to 5 x 10^1^ copies of DNA/μL. This equated to approximately 10^6^ to 10^0^ copies of DNA/reaction. A 96-well primer plate (IDT) with pre-mixed forward and reverse primers in a single well was diluted to 100 μM with the addition of 40 μL of NFW to each well. A total of 2.5 μL of the primer mix (100 μM) was added to 97.5 μL of NFW for a final concentration of 2.5 μM. The same primers as qPCR were used for nl-qPCR (Table S1). A master mix consisting of 473 μL SybrGreen 2x Master Mix (Thermo Fisher Scientific) and 284 μL of NFW was prepared. In a 384-well plate (Takara Bio USA), 14.3 μL of master mix and 3.6 μL of 2.5 μM primer mix were added into each well. Two wells were prepared for each primer as technical replicates. The assay plate was subsequently sealed, centrifuged at 1000 rpm for 1 min., and stored in −80 °C. A master mix consisting of 2472 μL SybrGreen 2x

Master Mix (Thermo Fisher Scientific) and 495 μL of nuclease free water was prepared. A 384-well plate (Takara Bio USA) was prepared with 7.5 μL of master mix and 5 μL of the tNA template from the sample. The plate was then sealed, centrifuged at 1000 rpm for 1 min., and stored at −80 °C. Plates were shipped overnight on dry ice to Washington State University along with the Takara Bio SmartChip^®^ MyDesign Kit. The Takara Bio SmartChip Real-Time PCR system is composed of the SmartChip MultiSample NanoDispenser (MSND) and the SmartChip Real-Time PCR Cycler. The MSND dispenses the samples and PCR assays into the nanowell chip. Each chip (5,184 wells) run on the SmartChip Real-Time PCR Cycler yields data which are pre-processed within the Takara system for subsequent post-processing and analysis that we performed. The cycler was run at 50 °C for 60 min., followed by 95 °C for 10 min., 40 cycles of 95 °C for 15 sec. and 60 °C for 1 min. A final melt curve was run at 65 °C for 10 seconds followed by 0.4 °C ramp per second to a final 97 °C. Data was collected using Wafergen qPCR software (Takara; v2.8.6.1). Ct values for samples that were between 35 and 40 were truncated to a final Ct = 35.

#### Analytic controls and standard curves

Serial dilutions of known concentrations of *Vc* (CFU/µL) and phage (PFU/µL; ICP1, ICP2, ICP3) were prepared for qPCR and nl-qPCR standard curve analysis. For qPCR*, Vc* and ICP1 were serially diluted from approximately 10^6^ to 10^-3^ CFU(PFU)/µL, ICP2 from approximately 10^3^ to 10^-6^ PFU/µL, and ICP3 from approximately 10^7^ to 10^-1^ PFU/µL. For nl-qPCR, *Vc* was serially diluted from approximately 10^6^ to 10^-4^ CFU/µL and phages were diluted from approximately 10^7^ to 10^-3^ PFU/µL. All dilutions were run in three technical replicates and averaged. Ct values for each target and approach were plotted against the log (concentration) (Figs S4, S5). The linear segment of the plots was analyzed by linear regression for a line of best fit to calculate the relationship between Ct value and true biologic target concentration. The positivity threshold was set at the point where linearity of Ct values was lost among the serial dilutions (Fig S4, S5).

### Statistical analysis

#### Data aggregation, cleaning and exclusions

Samples that were negative for the *16S* rDNA primer either by qPCR or nl-qPCR were designated as ‘run failures’ likely because of a failed reaction or the presence of a inhibitors. Run failure samples were removed from subsequent analyses for the respective assays. Analyses including PCR data excluded those samples missing PCR results for the *Vc* target. Samples missing self-reported antibiotic exposure (Dhaka site) were removed from machine learning analyses.

#### Alignment analysis

To evaluate the qualitative alignment across all molecular diagnostics, Fleiss’s kappa was computed using the irr package (v0.84.1) in R (25). Euler plots were used to visualize alignment of diagnostics for a given target. Cohen’s kappa was used to compare two-way qualitative alignment for a given target (PCR vs qPCR, qPCR vs nl-qPCR, and PCR vs nl-qPCR) (25).

#### Comparison of quantitative molecular data

To compare target quantifications (Ct values) by qPCR and nl-qPCR, we conducted a paired Mann-Whitney U test (Wilcoxon signed rank test) with continuity correction for each target using the stats package (v4.5.1) in R (26). The test was conducted on all samples assayed by both qPCR and nl-qPCR, as well as the subset of samples that were positive for the given target by both assays.

#### Ranking factors associated with Vc diagnostic malalignment

Clinical (self-reported antibiotic exposure, dehydration status, emesis, and duration of diarrhea), sociodemographic (age, sex, sample site), and microbiological (ICP1 and ICP3 quantity) factors were analyzed via machine learning (ML) analysis to predict *Vc* assay alignment among cholera patients (*Vc*-positive by at least one assay). In the initial preparatory analysis, all available factors (*n*=18) were ranked using a random forest (RF) algorithm. Microbiologic features for phage (*n*=2) were selected based on the RF ranking. Clinical (*n=*4) and sociodemographic (*n*=3) features were selected based on prior knowledge of clinical relevance irrespective of the initial ranking (8, 11, 27). Subsequently, five classification ML models (random forest, eXtreme Gradient Boosting, categorical boosting, logistic regression, and k-nearest neighbors) were developed based on a finalized set of features (*n*=9) (28–33). The top model was selected by comparing the receiver operating characteristic-area under curve (ROC-AUC) performance metric across all models for the test data. SHapley Additive exPlanation (SHAP) values (34) were utilized to rank and explain how top features contributed to the predicted result for *Vc*-diagnostic alignment or malalignment. Additional detailed methodology for data preprocessing, feature importance analysis, and ML model development is provided in the Supplementary Methods.

#### Cost comparison

A cost comparison analysis was conducted based on publicly available supplier price information (2026 values). Cost of reagents were calculated by averaging values across three suppliers (excluding products proprietary to single suppliers), assuming they were representative of typical costs in 2026. Cost attribution for PCR included both the gel matrix and reaction mixture. The cost of machinery was excluded from analysis, assuming in-kind machinery access.

**FIG 1.**
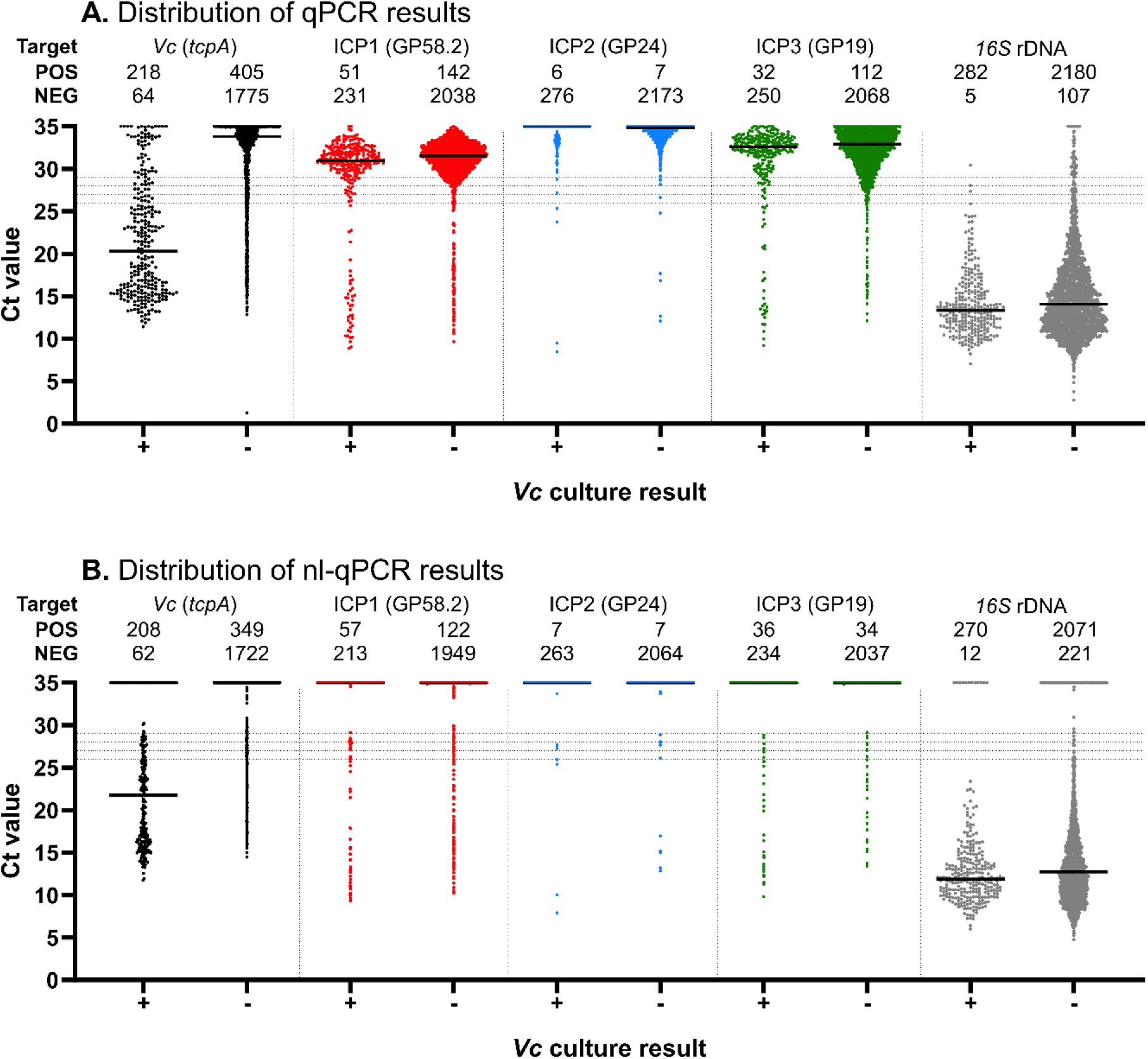
Comparison of qPCR (A) and nl-qPCR (B) between samples that were *Vc* culture positive (‘+’) or culture negative (‘-‘). Dashed lines represent Ct values of 26, 27, 28, and 29 of which < 28 was used as the threshold to define positivity; the threshold for *16S* rDNA positivity was set to <26. Enumerations for positive (‘POS’) and negative (‘NEG’) results are shown for all targets; enumerations for *Vc* and phage targets do not include run failures (*16S* rDNA ≥ 26).

## RESULTS

### Patient characteristics

As described in the parent study, a total of 4,975 patients were enrolled (14); 2,574 stool samples were collected. Among samples available for analysis, a total of 533 (20.7%) were from Habiganj, 281 (10.9%) from Naogaon, 240 (9.3%) from Patuakhali, 342 (13.3%) from Chattogram, 190 (7.4%) from Cumilla, 594 (23.1%) from Kusthia, and 394 (15.3%) from Dhaka (Table 1). 1,258 (48.9%) participants were male. The median age was 25 years (IQR: 1.44-40). Clinically, 1,524 (59.2%) participants presented with emesis, and the median duration of diarrhea was 48 hours (IQR: 24-72). A total of 440 (17.1%) patients had mild dehydration, 1,338 (52%) had moderate dehydration, and 796 (30.9%) had severe dehydration. A total of 1,089 of 2,180 (50%) responded “yes” to prior antibiotic exposure; self-reported antibiotic exposure data was unavailable for Dhaka.

**TABLE 1.** Patient characteristics with stratification for *Vc* assay alignment.

| Characteristic | All samples<br>N (%) | Molecular assay alignment for<br>Vc |  |
| --- | --- | --- | --- |
|  |  | Yes<br>N (%)<sup>a</sup> | No<br>N (%)<sup>b</sup> |
| Total | 2574 (100%) | 315 (100%) | 240 (100%) |
| Age (Years; Median [IQR]) | 25 [1-40] | 28 [12-43] | 25 [2-39] |
| Sex: Male | 1258 (48.9%) | 150 (47.6%) | 111 (46.3%) |
| Dehydration status |  |  |  |
| Mild | 440 (17.1%) | 51 (16.2%) | 56 (23.3%) |
| Moderate | 1338 (52.0%) | 167 (53.0%) | 123 (51.3%) |
| Severe | 796 (30.9%) | 97 (30.8%) | 61 (25.4%) |
| Area code |  |  |  |
| Habiganj (#2) | 533 (20.7%) | 47 (14.9%) | 51 (21.3%) |
| Naogaon (#4) | 281 (10.9%) | 36 (11.4%) | 23 (9.6%) |
| Patuakhali (#05) | 240 (9.3%) | 70 (22.2%) | 33 (13.8%) |
| Chattogram (#10) | 342 (13.3%) | 76 (24.1%) | 67 (27.9%) |
| Cumilla (#15) | 190 (7.4%) | 22 (7.0%) | 14 (5.8%) |
| Kusthia (#17) | 594 (23.1%) | 64 (20.3%) | 52 (21.7%) |
| Dhaka (#23) | 394 (15.3%) | <sup>-d</sup> | <sup>-d</sup> |
| Duration of diarrhoea<br>(Hours; Median [IQR]) | 48 [24-72] | 48 [24-72] | 48 [24-48] |
| Vomiting | 1524 (59.2%) | 227 (72.1%) | 135 (56.3%) |
| Self-reported antibiotic exposure | 2180 (84.7%) | 315 (100%) | 240 (100%) |
| Yes | 1089 (50.0%)<sup>c</sup> | 176 (55.9%) | 140 (58.3%) |
| No | 1091 (50.0%)<sup>c</sup> | 139 (44.1%) | 100 (41.7%) |

<sup>a</sup> Samples positive for Vc by PCR, qPCR, and nl-qPCR (i.e. positive alignment) that were included in ML analyses
<sup>b</sup> Samples positive for Vc by at least one but fewer than three molecular diagnostics that were included in ML analyses
<sup>c</sup> Percent reported from the total number of patients with data for self-reported antibiotic
<sup>d</sup> Samples missing self-reported antibiotic exposure (Dhaka site) removed from ML analyses

### *Vc* detection and diagnostic alignment

Standard curves were generated for qPCR (Fig S4) and nl-qPCR (Fig S5) which set positivity thresholds at 7.3×10^5^ and 7.5×10^5^ CFU/mL, respectively. All 2,574 samples were tested for *Vc* culture result. A total of 2,588 samples were analyzed by PCR; 644 samples were run in duplicate resulting in 72 (11.2%) conflicts that were resolved with a third replicate. qPCR was performed on 2,462 samples; 112 samples were negative for *16S* rDNA and removed from subsequent analysis. nl-qPCR was performed on 2,341 samples; 233 were *16S* rDNA negative and removed from subsequent analysis. The sample sizes after exclusions for qPCR and nl-qPCR are equal for all targets (*Vc*, ICP1/2/3). Among all samples, *Vc* was detected by culture in 11% (*n=*287) of samples. Among the samples for which all three assays were performed for a given target, *Vc* was detected by PCR in 33% (*n=*850) of samples, qPCR in 25% (*n*=623) of samples, and nl-qPCR in 24% (*n*=557) of samples (Table S2). There was strong three-way assay alignment (PCR, qPCR, nl-qPCR) for *Vc* (κ=0.785) (Table S3; Fig 2). Among assay-specific *Vc*-positive samples, the median *Vc*-positive qPCR Ct value was 21.0 (IQR: 16.9-25.3) and nl-qPCR Ct value was 21.0 (IQR: 17.5-25.4) (Table S2). Despite similar medians, there was a statistically significant difference in qPCR and nl-qPCR Ct values among all and positive-only samples by paired the Mann-Whitney U test (*P* < 0.001) (Table S4; Fig 3).

**FIG 2.**
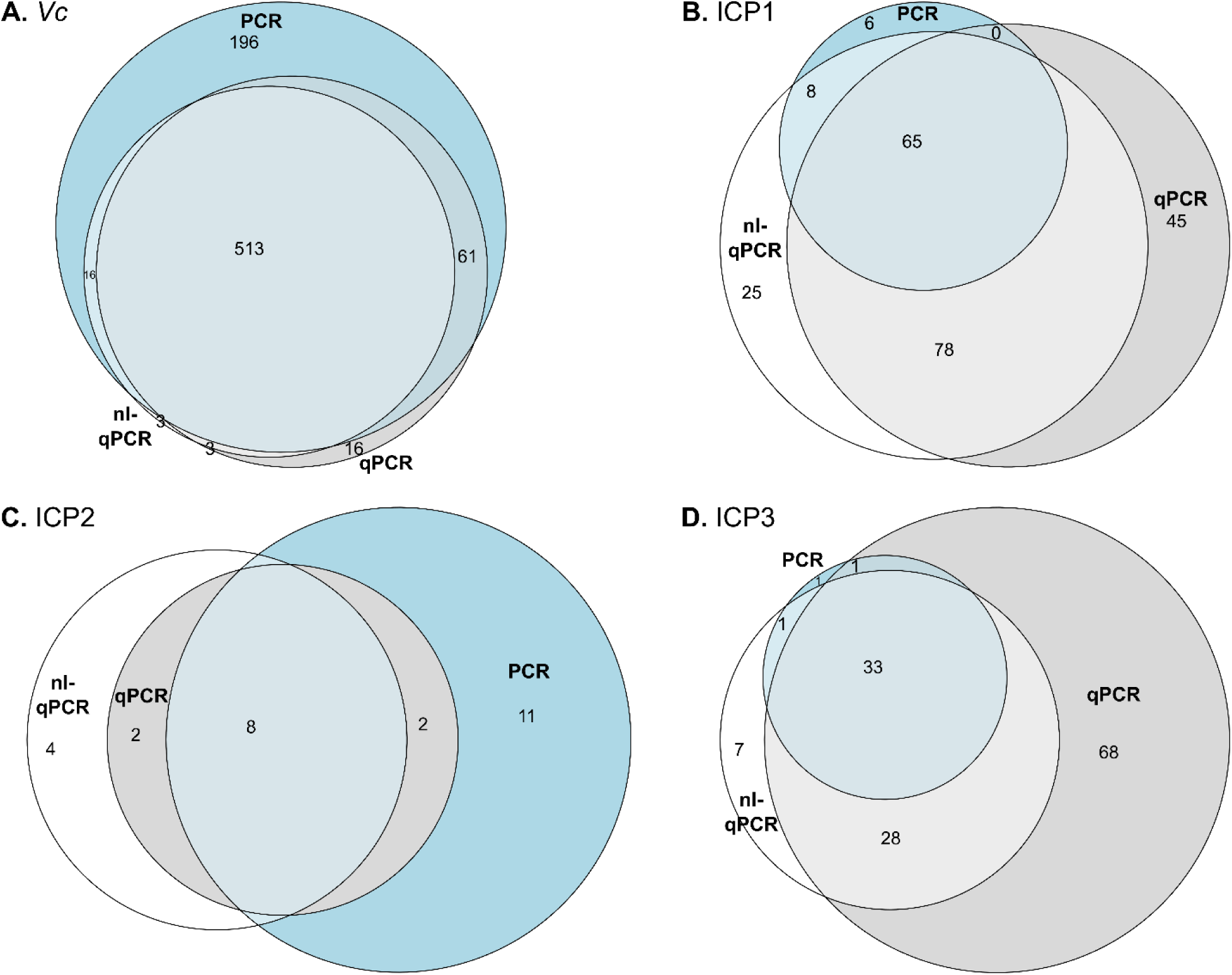
Alignment of positive results for target detection by PCR (blue), qPCR (grey), and nl-qPCR (white). Targets are *Vc* (**A**), ICP1 (**B**), ICP2 (**C**), and ICP3 (**D**). Results are displayed proportionally as a Euler diagram.

**FIG 3.**
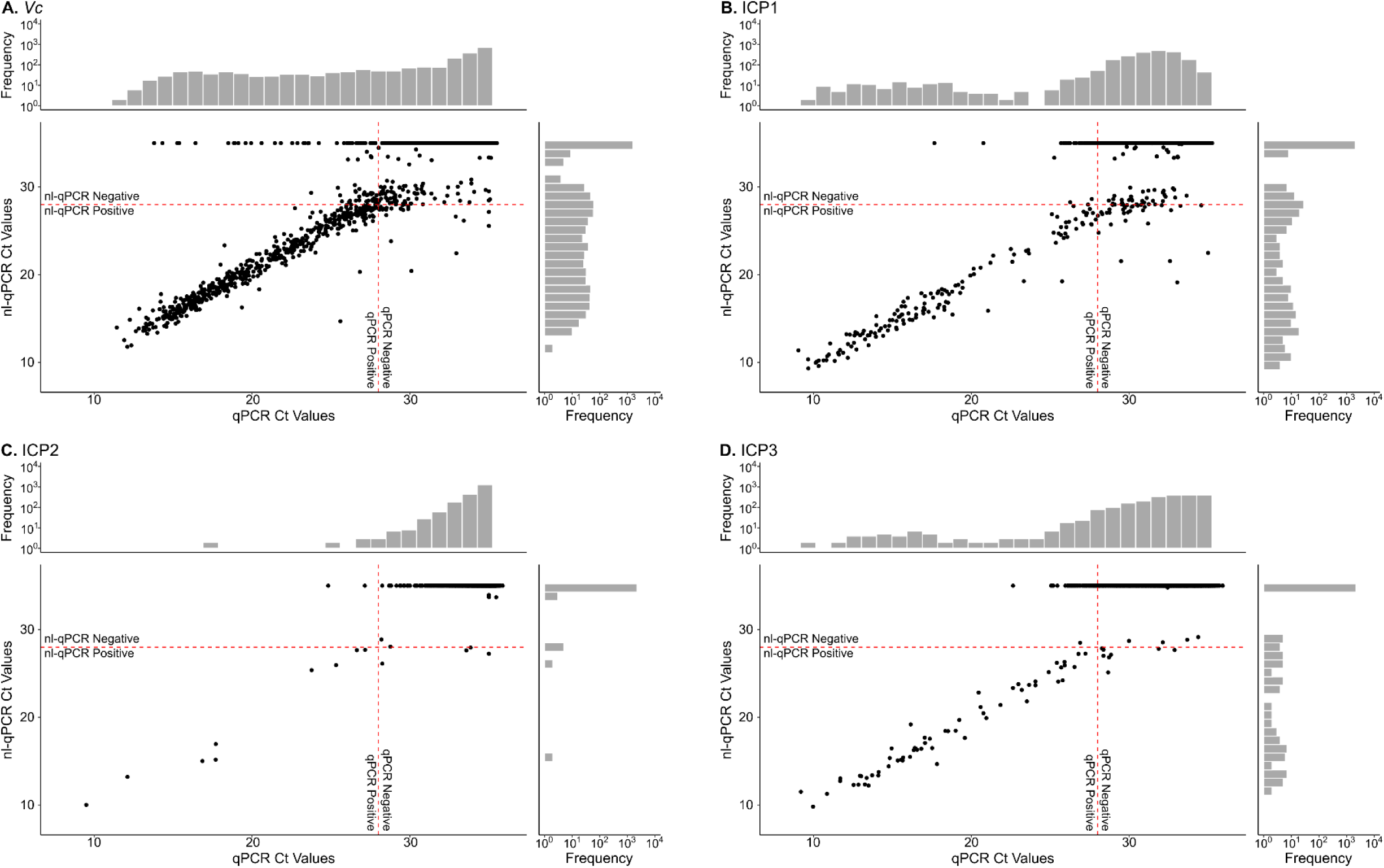
Comparison of qPCR and nl-qPCR Ct assay results. Distributions for (**A**) *Vc*, (**B**) ICP1, (**C**) ICP2, and (**D**) ICP3. Scatterplots show corresponding qPCR Ct values on the x-axis and nl-qPCR Ct values on the y-axis for each sample. Along the axes are histograms of the distribution of Ct values by diagnostic type. Dotted red lines indicate the cutoff for positivity (Ct<28); lower Ct values represent higher target abundance, and higher Ct values represent lower target abundance.

### ICP1 detection and diagnostic alignment

Standard curves were generated for qPCR (Fig S4) and nl-qPCR (Fig S5) which set positivity thresholds for ICP1 at 1.7×10^3^ and 2.0×10^3^ PFU/mL, respectively. ICP1 was detected by PCR in 4% (*n*=89), qPCR in 8% (*n*=193), and nl-qPCR in 8% (*n*=179) of samples (Table S2). There was moderate three-way assay alignment (κ=0.609) (Table S3; Fig 2). For ICP1-positive samples, the median qPCR Ct value was 18.5 (IQR: 14.8-26.3) and nl-qPCR Ct value was 17.9 (IQR: 14.0-25.7) (Table S2). Among all and positive-only samples, statistically significant differences were observed between qPCR and nl-qPCR Ct values (*P* < 0.001) (Table S4; Fig 3).

### ICP2 detection and diagnostic alignment

Standard curves were generated for qPCR (Fig S4) and nl-qPCR (Fig S5) which set positivity thresholds for ICP2 at 9.3×10^3^ and 1.6×10^3^ PFU/mL, respectively. ICP2 was detected by PCR in 1% (*n*=26), qPCR in 0.5% (*n*=13), and nl-qPCR in 0.6% (*n*=14) of samples (Table S2). There was moderate three-way assay alignment for ICP2 (κ=0.593) (Table S3; Fig 2). For ICP2-positive samples, the median qPCR Ct value was 17.7 (IQR: 12.7-25.3) and nl-qPCR Ct value was 25.7 (IQR: 15.1-27.5) (Table S2). There were statistically significant differences in Ct values between qPCR and nl-qPCR for all samples (*P* < 0.001) but not for positive-only samples (*P* = 1) (Table S4; Fig 3).

### ICP3 detection and diagnostic alignment

Standard curves were generated for qPCR (Fig S4) and nl-qPCR (Fig S5) which set positivity thresholds for ICP3 at 3.0×10^5^ and 2.2×10^5^ PFU/mL, respectively. ICP3 was detected by PCR in 2% (*n*=53), qPCR in 6% (*n*=144), and nl-qPCR in 3% (*n*=70) of samples (Table S2). There was moderate three-way assay alignment for ICP3 (κ=0.533) (Table S3; Fig 2). For ICP3-positive samples, the median qPCR Ct value was 26.0 (IQR: 18.2-27.5) and nl-qPCR Ct value was 18.1 (IQR: 14.8-24.1) (Table S2). There were statistically significant differences in Ct values between qPCR and nl-qPCR for all samples (*P* < 0.001) but not for positive-only samples (*P* = 0.38) (Table S4; Fig 3).

### Relative importance of phage predation on *Vc*-diagnostic alignment

Among 2,574 samples, 2,245 were assayed for *Vc* by PCR, qPCR, and nl-qPCR. Cholera patients were defined as *Vc*-positive by at least one assay (*n*=808). Samples missing self-reported antibiotic exposure data were excluded from downstream analyses (*n*=253). Effectors of *Vc*-diagnostic alignment were identified and ranked with ML classification models among a final set of *n*=555 patients. Preliminary feature ranking by a RF model found that phage Ct values by qPCR were more important as a predictive variable than phage Ct values by nl-qPCR or absolute phage detection (Fig S6). The finalized list of features included phage (ICP1 and ICP3) quantification (Ct values) by qPCR, age, diarrheal duration, emesis, sex, area code, dehydration status and self-reported antibiotic exposure; features for *Vc*-aligned and *Vc*-mal-aligned samples are described in Tables 1, S2 and S6. After comparison of ROC-AUC on test data across ML models, the random forest model was the highest performing with a ROC-AUC of 0.70.

Additional performance metrics for all five ML models tested for cross-validation and test data are provided (Table S5). SHAP analysis indicated that phage were the first (ICP1) and third (ICP3) most impactful predictors of *Vc-*diagnostic alignment (Fig 4). Age (second), emesis (fourth), diarrheal duration (fifth) and self-reported antibiotic (sixth) were also highly ranked.

**FIG 4.**
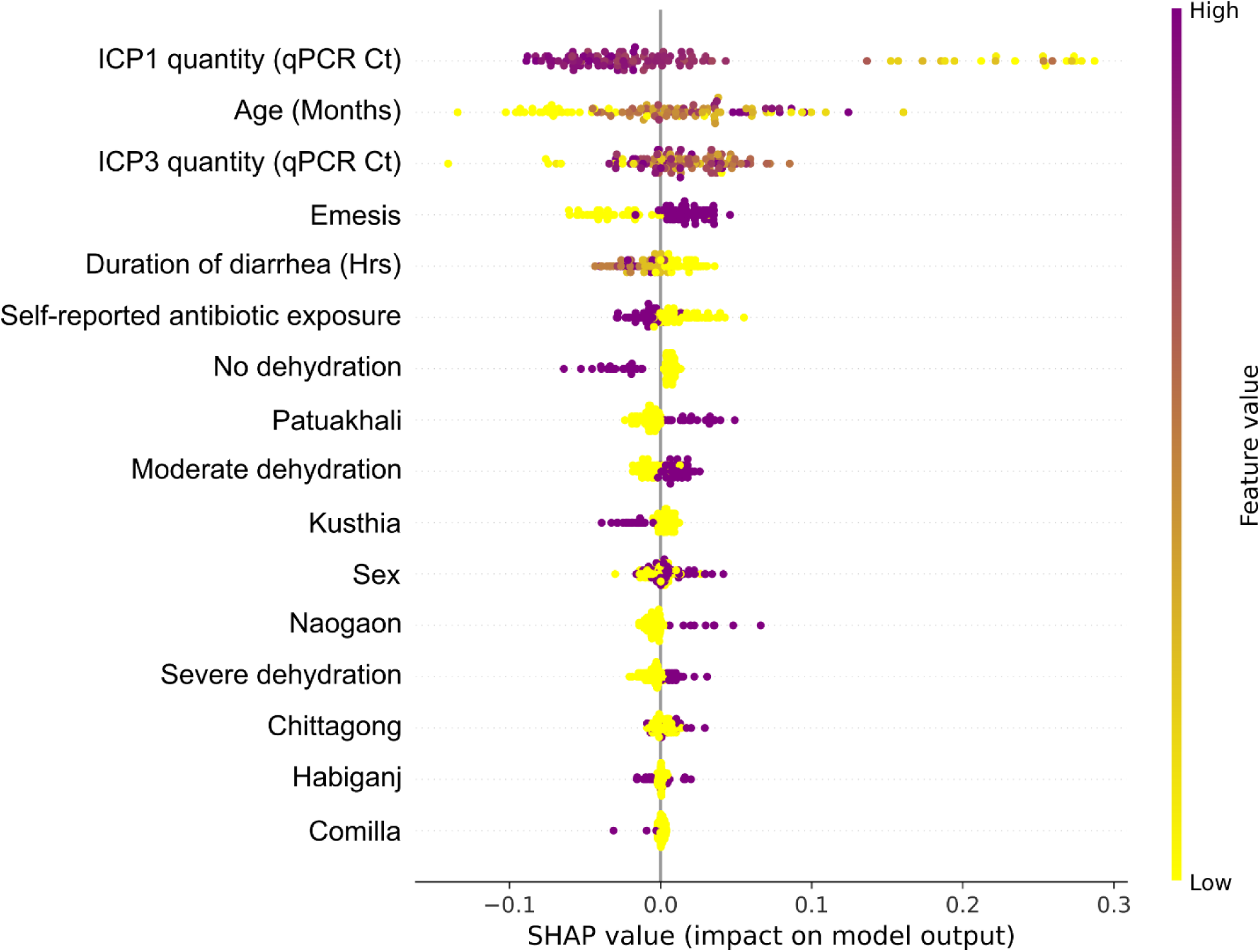
Relative importance of phage as a determinant of *Vc* molecular diagnostic alignment. SHAP beeswarm plot based on the final model with the highest ROC-AUC performance on test data. The plot shows the relationship between raw feature value and relative impact on model prediction of alignment (rightward positive SHAP) or malalignment (leftward negative SHAP) for each sample in the test data (represented by a circle for each feature). Yellow indicates a low feature values for continuous variables and negative for binary variables. Purple represents a high feature value and positive for binary variables. qPCR Ct values are low when the target is in high abundance and high when the target is at low abundance.

### Cost comparison analysis

A cost comparison analysis based on the cost of reagents was conducted, assuming no-cost (in-kind) access to machinery The cost per reaction (CPR) averaged across three suppliers found PCR was most expensive ($1.58 CPR) compared to qPCR ($1.19 CPR) and nl-qPCR ($0.20 CPR). The high cost of PCR was attributed to the cost of gel-matrix. Given the comparatively low, nanofluidic volumes of reagents required in nl-qPCR, the majority of nl-qPCR costs were attributed to the SmartChip MyDesign Kit (Takara Bio USA, Inc.) which is only available through Takara.

## DISCUSSION

PCR-based molecular assays represent a fast and accurate method to detect *Vc* and diagnose cholera. However, deciding which molecular diagnostic to deploy is challenging, especially in resource limited settings. This study focused on the alignment of results from different molecular assays for the detection of *Vc* and associated phages. We found the degree of alignment of diagnostic results was highest for the detection of *Vc* compared to phages. However, there remained samples in which *Vc* assays did not align and phage ranked as the first (ICP1) and third (ICP1) factors impacting *Vc* assay alignment. Taken together, these findings provide actionable insight into the strengths and limitations of different molecular assays to detect *Vc* and associated phages as stakeholders develop molecular diagnostic panels for resource-limited settings.

The first objective of this study was to determine the degree to which molecular assays aligned. To address this objective the technical performance of target detection was assessed through a series of standard curves using biologic targets; synthetic targets were used as internal controls for nl-qPCR. Serial dilutions of the targets produced linear results across biologically meaningful concentrations. The threshold to differentiate samples as positive or negative was based on the combined results from standard curves, Ct values for in-plate positive and negative controls (no-template), as well as *16S* rDNA values to identify run failures by inappropriately low bacterial detection. While we set the thresholds at 28 for this study, Ct thresholds will vary between studies. In this context, assay alignment among clinical samples for *Vc* was higher than alignment for detection of the phages. *Vc* detection rates were highest by PCR compared to qPCR and nl-qPCR. ICP1 detection rates were highest by qPCR. ICP2 detection rates were low which limited subsequent analysis. ICP3 detection rates were equivalent. When comparing target quantification among positive samples, Ct values were only statistically different for the ICP1 target. The mechanism for lower Ct values (more target) in the nl-qPCR reaction is unknown; one explanation may be a dilutional effect of reaction inhibitors from the stool matrix.

The second objective was to identify factors that impact the alignment of assays to detect *Vc*. We conducted an exploratory analysis using machine learning (ML) and model development. The analysis focused on phage because prior studies found phage impact disease severity as well as an array of diagnostics (e.g. rapid diagnostic tests, culture, direct microscopy and qPCR) (8, 18, 35). After an initial feature ranking and selection process, the final analyses revealed that ICP1 and ICP3 ranked as the first and third most important factors that impacted the alignment of *Vc* molecular assays.

Their effects are likely due to simple target elimination such as nucleic acid degradation by phage-encoded nucleases as well as more complicated predator-prey dynamics. For example, we have previously shown that the relative abundance of phage (predator) in patients shares a quadratic relationship with the relative abundance of *Vc* (prey) in the human host (18), which is impacted by multiple factors, including antibiotic exposure. In this study, higher ICP1 abundance (lower Ct value) was associated with alignment, however higher ICP3 abundance was associated with malalignment. These findings support the proposal that virulent phages can compromise diagnostic performance and therefore their detection should be considered as part of diagnostic panels.

The third objective was to conduct a cost comparison analysis. The analysis is important because the biological advantages and disadvantages of a given assay are relative to the associated cost of each assay. In the analysis we assumed no-cost access to machinery, which is reasonable given large investments in global health infrastructure over the past ten years. We found PCR was the most expensive method which was due largely to the cost of gel matrix. Logistically, the physical manipulation of large-format PCR gels is also difficult, and gel analysis is vulnerable to subjective interpretation. We found qPCR ranked second in cost. The least costly was nl-qPCR despite posing the greatest barriers for accessing machinery. Taken together, qPCR represents a reasonable balance between target detection, cost, and accessibility, however the other methods remain advantageous in select contexts.

These findings should be considered within the context of their limitations. First, the primers for *Vc*, ICP1, and ICP3 differed between PCR and qPCR/nl-qPCR which may have impacted differences in assay alignment. Second, our approach favored the robustness of technical replicates for a single genetic target for *Vc* over having primers-sets for multiple genetic targets (e.g. cholera toxin, LPS, outer membrane proteins) which is a common approach of commercialized kits. This leaves the possibility that *Vc* detected with *tcpA* primers alone will not differentiate less common non-O1 *Vc* and will result in a false negative if there is a rare *tcpA* variant. Third, the alignment analyses did not account for objectively detected (mass spectrometry) antibiotic exposures which may be a determinant for target detection rates and alignment (11). Lastly, the analysis to identify and rank factors that impact *Vc* assay alignment was exploratory and limited by small sample sizes and low accuracy. Future models will need larger training datasets to increase accuracy. Despite these limitations, the findings add critical information to the larger body of literature on cholera diagnostics and offer important insights into developing and scaling molecular methods to identify cholera cases.

## CONCLUSION

This study found differences between three PCR-based assays to detect *Vc* and associated phages. Phages ranked as the first and third most important factors impacting the alignment of assays to detect *Vc*. While each assay has limitations, qPCR offers a reasonable compromise between cost and target detection for scaled clinical and research applications. Additionally, the impact of phage on *Vc* assay alignment supports the rationale to include phage detection as a proxy for *Vc* detection in molecular cholera diagnostic panels. These findings are important and relevant to less tractable bacterial diseases that face similar diagnostic challenges.

## Data Availability

Data are available upon request.

## Code Availability

Code available upon request.

## Acknowledgements

We thank the patients for their participation in the parent study. We thank our diligent clinical and laboratory team members. We are grateful to the Institute of Epidemiology, Disease Control and Research (IEDCR), Ministry of Health and Family Welfare, and Government of Bangladesh who collaborated on the parent clinical study. We are also grateful to the Governments of Bangladesh and Canada for providing unrestricted support to the icddr,b. We are grateful to B. Johnson, T. Linn, N. Rushing and K. Berquist for their administrative guidance at the University of Florida.

## Author Contributions

Conceptualization: MAS, FQ, AIK, EJN Implementation of clinical study: MAS, FQ, AIK, EJN

Investigation and laboratory procedures: MAS, ETC, ACM, KI, MIUK, MTRB, YB, EF, AV, LB, DGB, DJP, JAG, EJN

Statistical analyses and visualizations: IS, MM, KM, ZL, EJN Funding acquisition: DTL, EJN

Supervision: ZL, JAG, FQ, AIK, EJN Writing – original draft: IS, MM, EJN

Writing – review & editing: IS, MM, MAS, ACM, DGB, DJP, KM, ZL, FQ, AIK, EJN

