## Supplementary Methods for "Molecular Methods to Detect *Vibrio cholerae* and Associated Bacteriophages among Diarrheal Patients in Bangladesh"

Cholera, diarrhoea, *Vibrio cholerae*, bacteriophage, phage, diagnostics, Bangladesh

**Enumerations**

Supplement:
- Methods: 675
- Figures: 6
- Tables: 7

References: 20

### SUPPLEMENTARY METHODS

**Machine learning (ML) analysis**. ML based methods were used to analyze the relative contribution of clinical, sociodemographic, and microbiologic factors on *Vc* diagnostic alignment. The sample distributions were insufficient to extend the approach to phage assays.

*Data preparation and preprocessing.* Samples negative for *Vc*-diagnostics across all three molecular approaches were removed from ML analyses. Ages reported in years were converted to months; diarrheal duration reported in days was converted to hours. Binary categorical variables were recorded as '0' for negative and '1' for positive. Variables with more than two categories (i.e., sample site and dehydration status) were structured to be compatible with the ML approach taken via OneHotEncoder (v1.6.1). Quantitative variables were normalized to 0 to 1 range using MinMaxScaler (v1.6.1). The output was *Vc*-diagnostic alignment, coded as '0' for mal-aligned or '1' for aligned (1). Variables (features) included in the ML analyses are described in Table S6; samples with missing data for these variables were removed from subsequent analyses. Feature selection was conducted both by ranking a total of 18 features in a random forest (RF) model (Fig S6) and based on prior evidence. Clinical features (*n*=4) included self-reported antibiotic exposure, dehydration status, emesis and duration of diarrhea (hrs). Sociodemographic features (*n*=3) include age, sex, and sample site. Microbiological features (*n*=10) included ICP1 detection (PCR, qPCR, and nl-qPCR), ICP3 detection (PCR, qPCR, and nl-qPCR), ICP1 quantity (qPCR Ct and nl-qPCR Ct), and ICP3 quantity (qPCR Ct and nl-qPCR Ct). A Bayesian optimized ten-fold cross validation (CV) model was utilized to rank features by feature importance (2). Clinical and sociodemographic features were selected based on prior knowledge of relevance (3–5), even when ranked lower by the RF model. Selection of microbiologic features was based on the feature importance ranking of the RF model. Phage quantification (Ct value) ranked higher than detection alone for both ICP1 and ICP3. Additionally, quantification by qPCR ranked higher than quantification by nl-qPCR. The final feature set (*n*=9) included clinical and sociodemographic features of interest along with phage (ICP1 and ICP3) quantification by qPCR for subsequent analyses.

*ML model pipeline development hyperparameter tuning.* Machine learning analyses were performed using NumPy (v2.0.2), Pandas (v2.2.2), catboost(v1.2.10), xgboost(v3.2.0) and scikit-learn (v1.6.1) libraries in Python (v3.12.13) (1, 6–9). The narrowed down list of features was analyzed using the random forest (RF), eXtreme Gradient Boosting (XGB), categorical boosting (Catboost), k-nearest neighbor (kNN), and logistic regression (LR) algorithms (1, 2, 10–13). The data were split into training (80%) and testing (20%) sets. Categorical features with more than two categories were one-hot encoded (except for Catboost) and numerical features were scaled from 0 to 1 range. Hyperparameters were tuned using Bayesian optimization and ten-fold cross validation (Table S7). All analyses related to ML development were conducted in Google Colab.

*Performance metrics.* Performance metrics measured include ‘balanced accuracy,’ sensitivity, specificity, positive predictive value (PPV), negative predictive value (NPV), F1 score, receiver operating characteristic - area under curve (ROC-AUC), and Matthews Correlation Coefficient (MCC) (1). Models were primarily compared based on their ROC-AUC scores for the ten-fold cross-validation and final test data.

*Analyzing features contribution to predicting Vc-diagnostic malalignment*. SHapley Additive exPlanations (SHAP) values were used to rank and analyze features for the "best" model selected by comparing ROC-AUC (14). SHAP values allow for determining the importance of features used in an ML model and understanding how each feature contributes to the individual predictions of the model using an approach rooted in game theory. SHAP values were also used to determine the relationship between the values of each feature and the predicted output.

*Limitations*

Initial feature selection was conducted on the entire dataset, potentially biasing the performance of the final RF model. However, RF ranking was only used to select microbiological features, the remaining were selected based on prior knowledge of clinical relevance. Additionally, balanced accuracy for testing data was moderate, limiting interpretations of feature ranking and applications for predicting *Vc*-diagnostic alignment. We hypothesize that the small sample size for both training and testing as well as the limited number of features may have contributed to this.

### SUPPLEMENTARY FIGURES

#### Figure S1

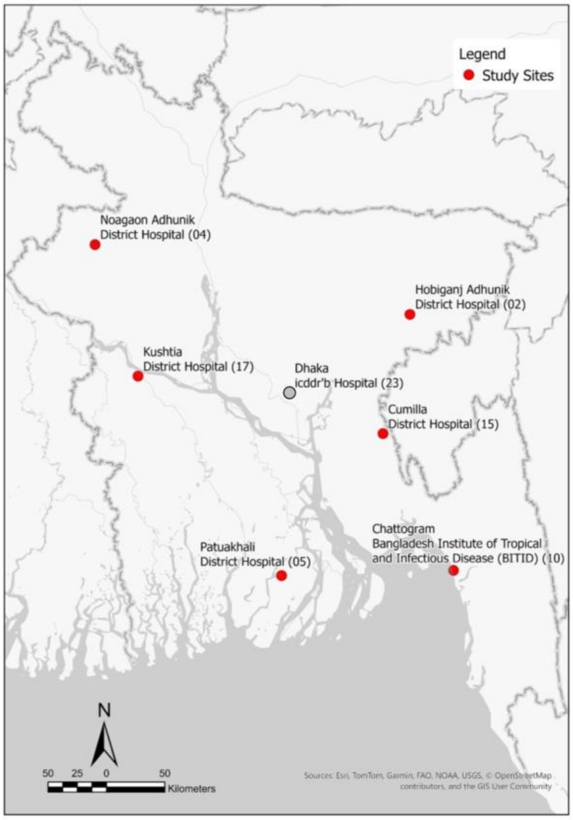

**FIG S1** Study sites map in Bangladesh (4). Numbers brackets indicate the site number and correlate with Table 1. Sites marked in red were included in machine learning analyses and sites marked in grey (Dhaka) were excluded.

**Figure S2**

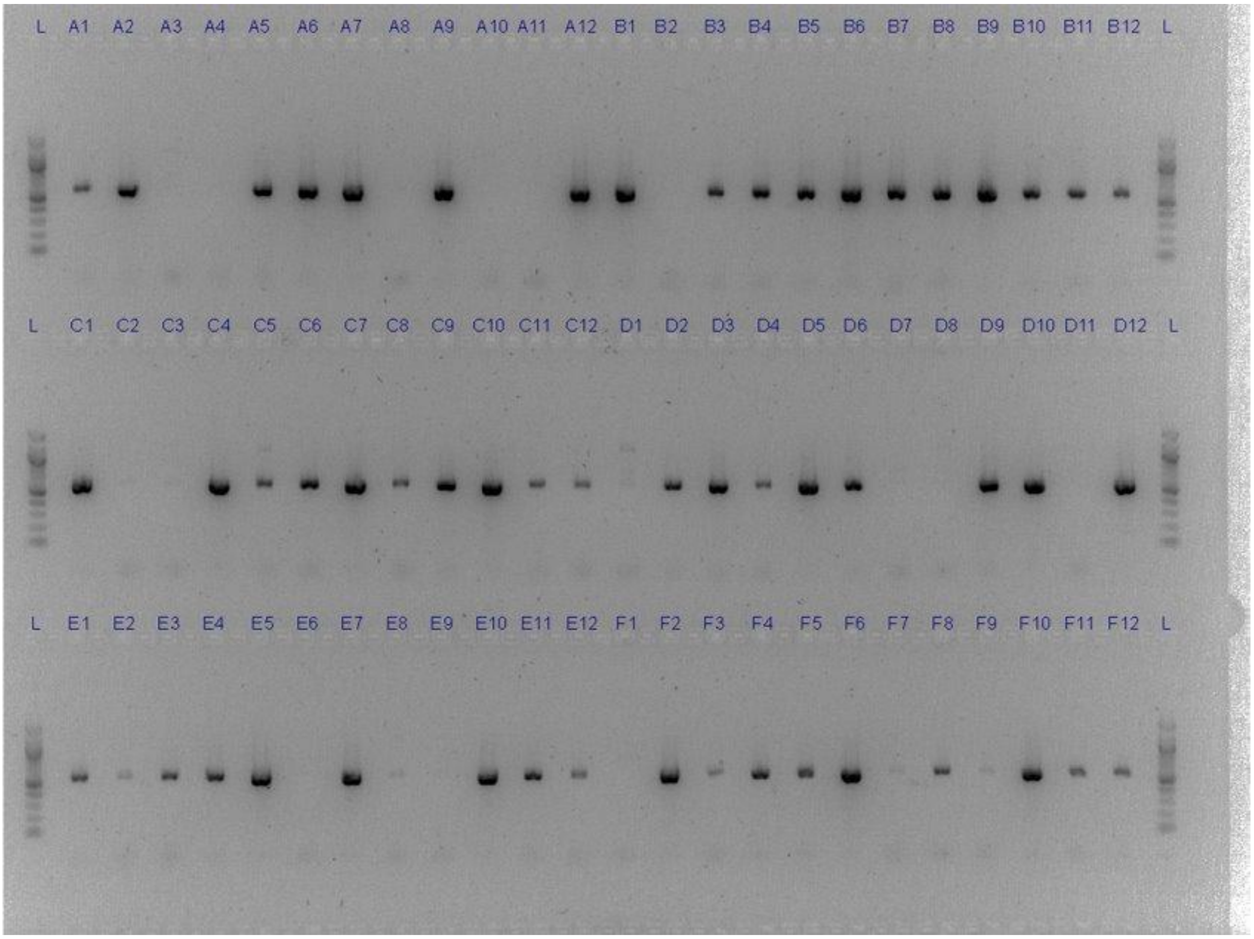

**FIG S2** Top of 96-well PCR gel for *Vc* (*ompW*) target with expected product size of 588 bases. 'L' indicates a ladder from 100 to 1000 bases. Original plate well locations of samples are labelled. B1 and E5 represent in-plate positive (*tcpA*) template controls; B2 and E6 represent negative controls.

**Figure S3**

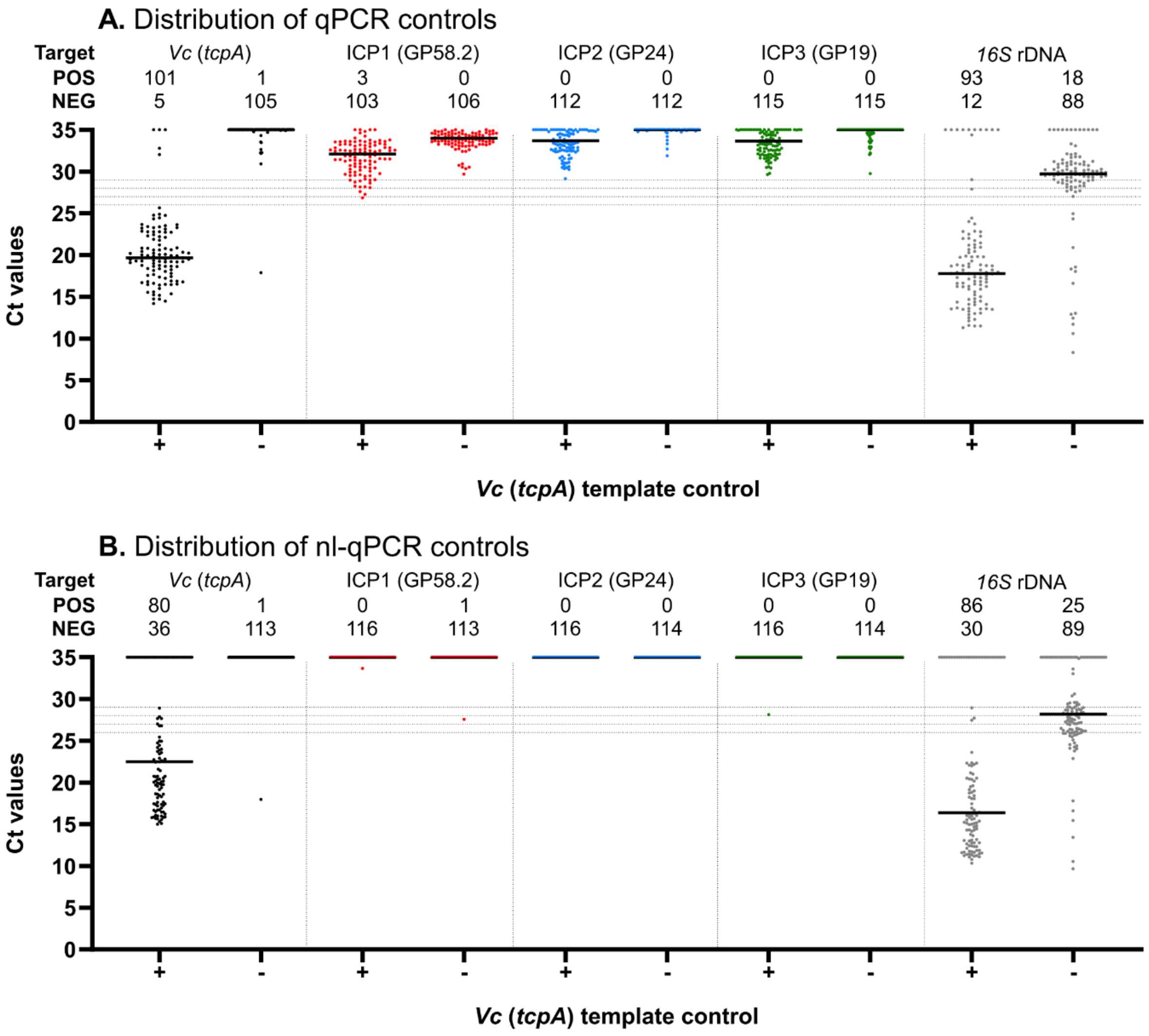

**FIG S3** Distribution of qPCR Ct values using in-plate *Vc* (*tcpA*) template controls and media negative controls. Dashed lines represent Ct values of 26, 27, 28, and 29 of which < 28 was used as the threshold to define positivity; the threshold for 16S rDNA positivity was set to <26. Enumerations for positive ('POS') and negative ('NEG') results are shown for all targets.

### Figure S4

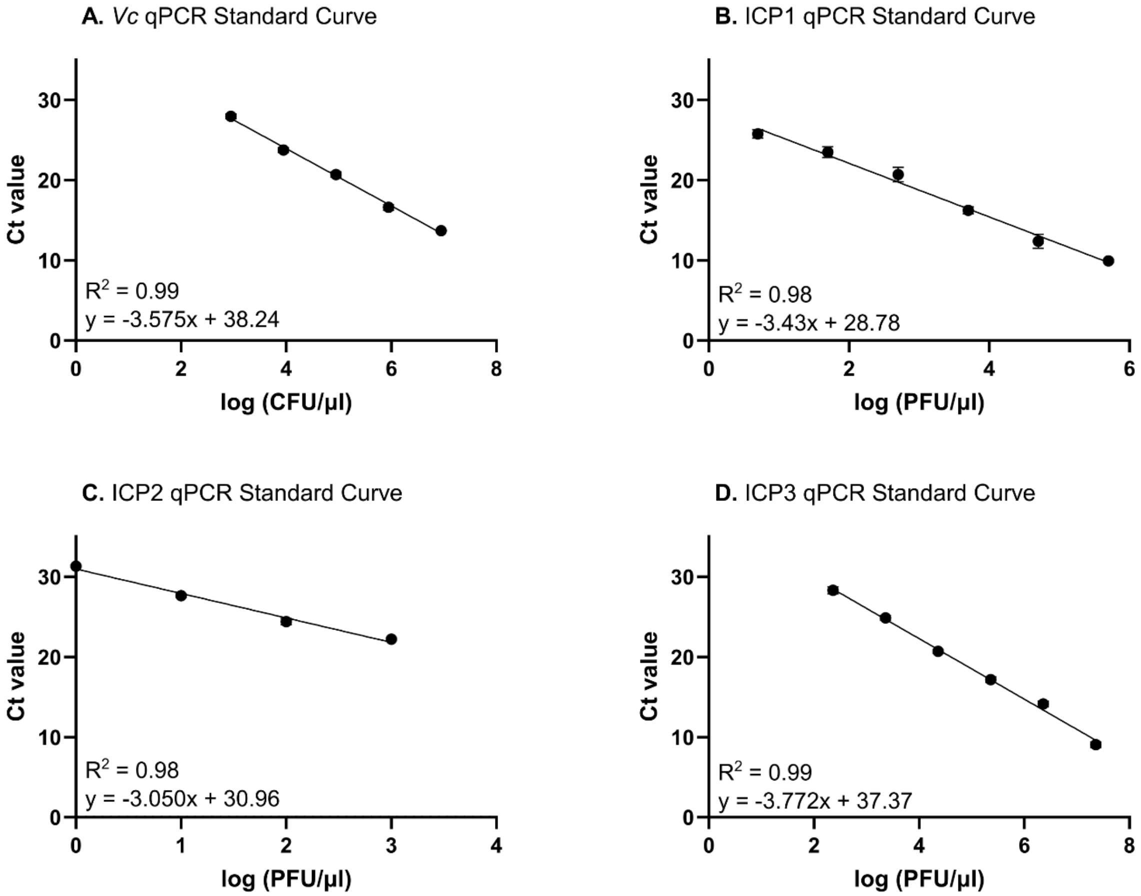

**FIG S4** Standard curves for absolute quantification of *Vc* (CFU/µL) and phage (PFU/µL) based on qPCR Ct values.

### Figure S5

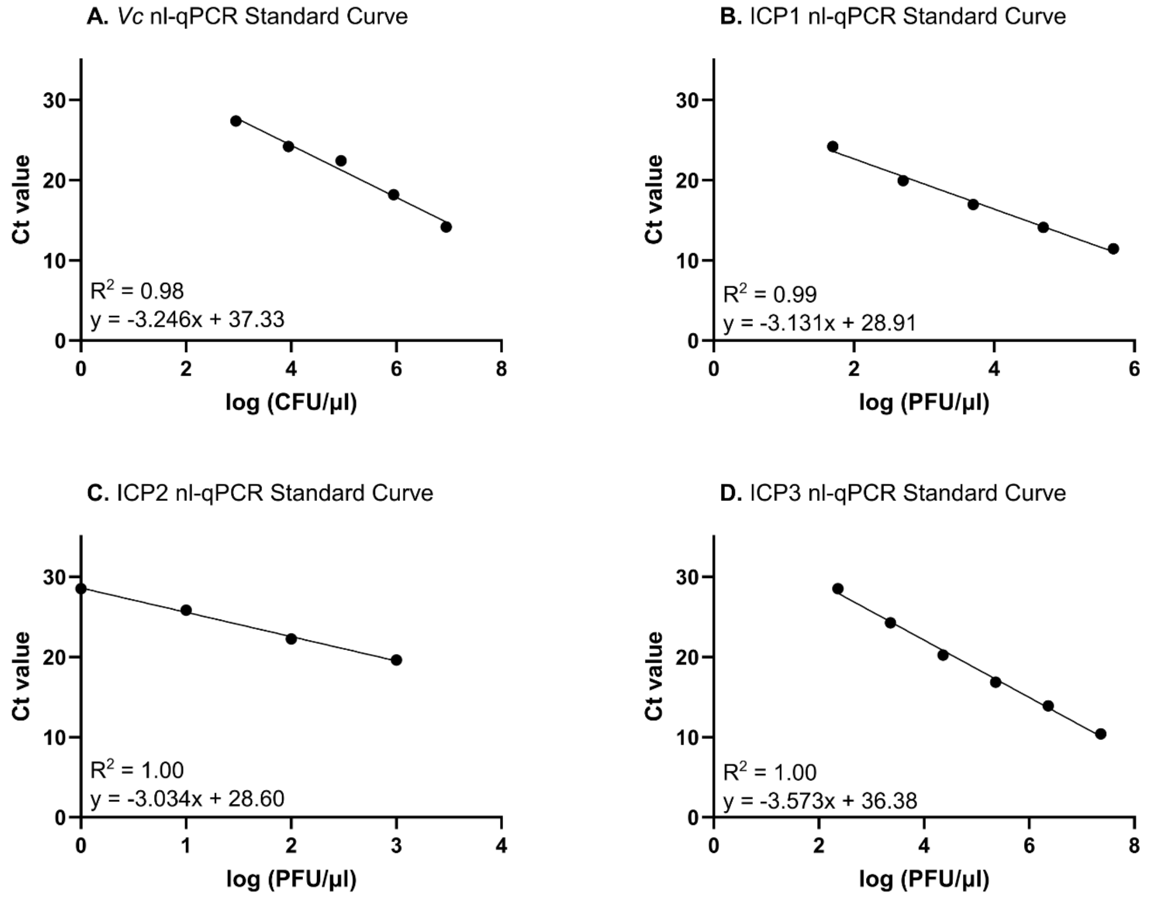

**FIG S5** Standard curves for absolute quantification of *Vc* (CFU/µL) and phage (PFU/µL) based on nl-qPCR Ct values.

**Figure S6**

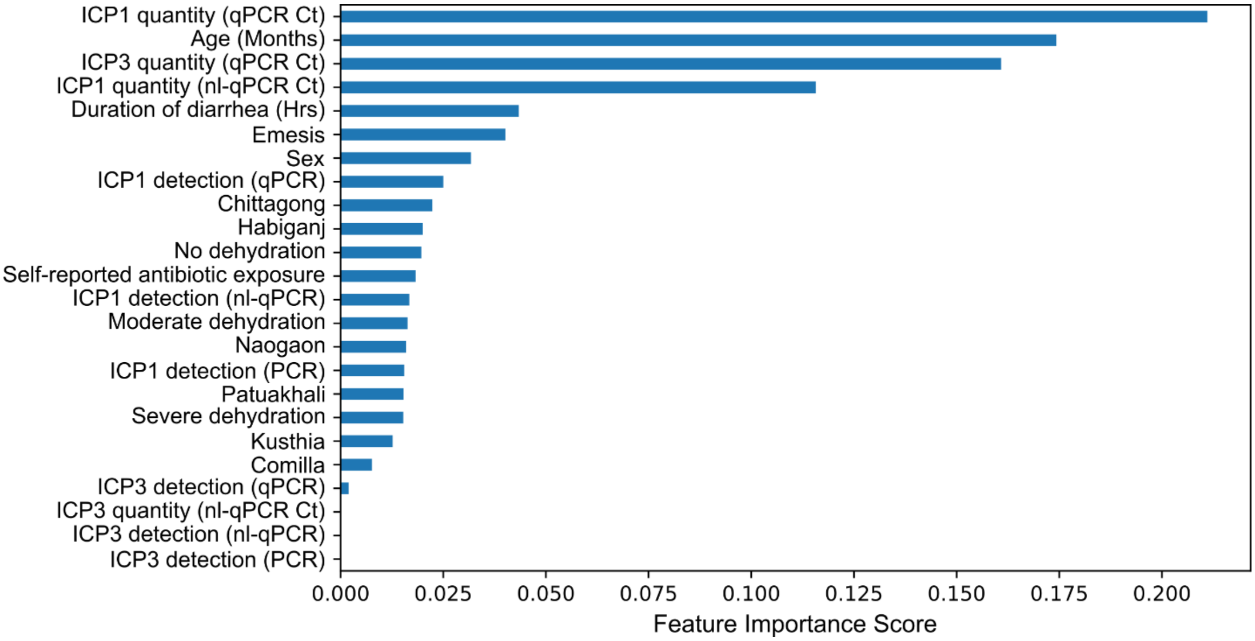

**FIG S6** Ranked list of feature importance in predictive random forest model for *Vc*-diagnostic malalignment

### SUPPLEMENTARY TABLES

**TABLE S1** Molecular diagnostic gene target primer sequences

| Target | Primer Name | Sequence 5'-3' | Reference |
|---|---|---|---|
| *Vc* | | | |
| *ompW* | ompW_F | CACCAAGAAGGTGACTTTATTGTG | (15) |
| | ompW_R | GAACTTATAACCACCCGCG | |
| *tcpA* | *tcpA set2*_F | ACACGATAAGAAAACCGGTCA | (16) |
| | *tcpA set2*_R | GCCTTGGTCATATTCTGCGA | |
| ICP1 | | | |
| GP58 | ICP1_gp58F | AACGCTGCTTTTCCTTTTGA | (17) |
| | ICP1_gp58R | CCCAGCATTGAGGACACTT | |
| | ICP1_gp58.2_F | CAAAGGCAGCAGGTAGGACA | (18) |
| | ICP1_gp58.2_R | CCCTTCAAGCCGTAGTTGGT | |
| ICP2 | | | |
| GP4 | ICP2_4F | CGCTAGTTCTGGCAGTGA GT | (19) |
| | ICP2_4R | TCCGTTCCAGTTCCAACAGG | |
| GP24 | ICP2_gp24_F | AGAAGTCGCAAACGGGGTAC | (19) |
| | ICP2_gp24_R | AACGTGGTTCTCGTGAGTGG | |
| ICP3 | | | |
| GP5 | ICP3_gp5F | ATTGTCGAGTGGGACAAAGG | (17) |
| | ICP3_gp5R | ACCAACTCGACGCATAGCTT | |
| GP19 | ICP3_gp19_F | AGACCAACGCCGACTGTTAG | (18) |
| | ICP3_gp19_R | CGATACCACGGAAAGCCTGT | |
| *16S* rDNA | Maeda_1048_1067_F | GTGSTGCAYGGYTGTCGTCA | (20) |
| | Maeda_1175_1194_R | ACGTCRTCCMCACCTTCCTC | |

**TABLE S2** Molecular detection and quantification of *Vc* and associated phage

| | All samples Pos./N (%) | *Vc* aligned Pos./N (%)[a] | *Vc* malaligned Pos./N (%)[b] |
|---|---|---|---|
| *Vc* | | | |
| PCR | 850/2558 (33.2) | 315/315 (100) | 220/240 (91.7) |
| qPCR | 623/2462 (25.3) | 315/315 (100) | 63/240 (26.3) |
| Median (IQR) – All[c] | 33.5 (27.8-34.9) | 21.59 (17.0-25.2) | 29.8 (27.9-33.0) |
| Median (IQR) – Positive[d] | 21.0 (16.9-25.3) | 21.59 (17.0-25.2) | 26.8 (25.5-27.6) |
| nl-qPCR | 557/2341 (23.8) | 315/315 (100) | 19/240 (7.9) |
| Median (IQR) – All[c] | 35.0 (28.4, 35.0) | 22.4 (18.0-25.9) | 35.0 (29.2-35.0) |
| Median (IQR) – Positive[d] | 21.0 (17.5-25.4) | 22.4 (18.0-25.9) | 27.4 (25.9-27.5) |
| ICP1 | | | |
| PCR | 89/2574 (3.5) | 23/315 (7.3) | 16/240 (6.7) |
| qPCR | 193/2462 (7.8) | 75/315 (23.8) | 23/240 (9.6) |
| Median (IQR) – All[c] | 31.5 (30.1-32.5) | 30.9 (28.7-32.0) | 31.4 (29.9-32.4) |
| Median (IQR) – Positive[d] | 18.5 (14.8-26.3) | 17.0 (14.3-23.5) | 19.2 (17.6-26.2) |
| nl-qPCR | 179/2341 (90.9) | 73/315 (23.2) | 23/240 (9.6) |
| Median (IQR) – All[c] | 35.0 (35.0-35.0) | 35.0 (33.5-35.0) | 35.0 (35.0-35.0) |
| Median (IQR) – Positive[d] | 17.9 (14.0-25.7) | 15.8 (13.6-20.7) | 19.3 (17.4-24.5) |
| ICP2 | | | |
| PCR | 26/2574 (1.0) | 6/315 (1.9) | 6/240 (2.5) |
| qPCR | 13/2462 (0.5) | 5/315 (1.6) | 3/240 (1.3) |
| Median (IQR) – All[c] | 34.9 (34.0-35.0) | 34.5 (33.6-35.0) | 34.8 (34.0-35.0) |
| Median (IQR) – Positive[d] | 17.7 (12.7-25.3) | 23.8 (9.5-25.3) | 17.7 (17.3-17.7) |
| nl-qPCR | 14/2341 (0.6) | 7/315 (2.2) | 3/240 (1.3) |
| Median (IQR) – All[c] | 35.0 (35.0-35.0) | 35.0 (35.0-35.0) | 35.0 (35.0-35.0) |
| Median (IQR) – Positive[d] | 25.7 (15.1-27.5) | 25.9 (17.7-26.7) | 15.2 (15.1-16.1) |
| ICP3 | | | |
| PCR | 53/2574 (2.1) | 1/315 (0.3) | 1/240 (0.4) |
| qPCR | 144/2462 (5.8) | 9/315 (2.9) | 9/240 (3.8) |
| Median (IQR) – All[c] | 32.8 (31.1-34.2) | 32.7 (31.6-33.9) | 32.7 (31.0-33.8) |
| Median (IQR) – Positive[d] | 26.0 (18.2-27.5) | 26.5 (22.6-27.3) | 26.5 (22.6-27.3) |
| nl -qPCR | 70/2341 (3.0) | 3/315 (1.0) | 3/240 (1.3) |
| Median (IQR) – All[c] | 35.0 (35.0-35.0) | 35.0 (35.0-35.0) | 35.0 (35.0-35.0) |
| Median (IQR) – Positive[d] | 18.1 (14.8-24.1) | 19.2 (18.8-21.3) | 18.5 (18.1-21.1) |

[a] Samples positive for *Vc* by PCR, qPCR, and nl-qPCR (i.e. positive alignment) that were included in ML analyses
[b] Samples positive for *Vc* by at least one but fewer than three molecular diagnostic that were included in ML analyses
[c] Median Ct values and IQR for all samples run with the associated diagnostic (qPCR or nl-qPCR) and target (ICP1/2/3/*Vc*)
[d] Median Ct values and IQR for samples positive (Ct < 28 for all targets) with the associated diagnostic (qPCR or nl-qPCR) and target (ICP1/2/3/*Vc*)

**TABLE S3** Qualitative comparison assay alignment

| | All (three-way)[a] | Two-way[a] | | |
|---|---|---|---|---|
| | | PCR + qPCR | qPCR + nl-qPCR | PCR + nl-qPCR |
| *Vc* | 0.785[b] | 0.753[b] | 0.886 | 0.722[b] |
| ICP1 | 0.609[b] | 0.474[b] | 0.767 | 0.556[b] |
| ICP2 | 0.593 | 0.553 | 0.768 | 0.417 |
| ICP3 | 0.533[b] | 0.437[b] | 0.597 | 0.641[b] |

[a] Fleiss' kappa was conducted for three-way comparisons and Cohen's kappa was conducted for two-way comparisons.

[b] Primer targets differed between molecular diagnostics. For *Vc*, PCR targeted *ompW* whereas qPCR and nl-qPCR targeted *tcpA*. For ICP1, PCR targeted a different region of the GP58 gene compared to qPCR and nl-qPCR. For ICP3, PCR targeted GP5 whereas qPCR and nl-qPCR targeted GP19.

**TABLE S4** Quantitative comparison of qPCR and nl-qPCR Ct values

| All samples | N | qPCR (Ct) Median (IQR) | nl-qPCR (Ct) Median (IQR) | Test statistic | *P* value |
|---|---|---|---|---|---|
| *Vc* | 2255 | 33.4 (27.5-34.8) | 35.0 (28.4, 31.4) | 177855 | <0.001 |
| ICP1 | 2255 | 31.5 (30.1-32.5) | 35.0 (35.0-35.0) | 78067 | <0.001 |
| ICP2 | 2255 | 34.8 (34.0-35.0) | 35.0 (35.0-35.0) | 226499 | <0.001 |
| ICP3 | 2255 | 32.7 (31.0-34.0) | 35.0 (35.0-35.0) | 46544 | <0.001 |
| Positive samples | N | qPCR (Ct) Median (IQR) | nl-qPCR (Ct) Median (IQR) | Test statistic | *P* value |
| *Vc* | 520 | 19.9 (16.7-24.2) | 20.6 (17.4-25.0) | 15850 | <0.001 |
| ICP1 | 143 | 17.0 (13.8-21.9) | 16.0 (13.4-21.1) | 7887 | <0.001 |
| ICP2 | 10 | 17.7 (13.3-24.9) | 16.1 (13.7-25.8) | 27 | 1 |
| ICP3 | 61 | 17.1 (14.8-23.0) | 17.1 (14.4-23.1) | 1068 | 0.38 |

[a] Paired Mann-Whitney U test for all samples processed via qPCR and nl-qPCR for each target (ICP1/2/3/*Vc*). Sample size (N), median, IQR, test statistic, and *P* value are reported. Median and IQR differ from Table S2 because samples are stratified to *16S* positive (Ct < 26) by both qPCR and nl-qPCR.
[b] Paired Mann-Whitney U test for samples positive by qPCR and nl-qPCR for each target (ICP1/2/3/*Vc*). Sample size (N), median, IQR, test statistic, and *P* value are reported. Median and IQR differ from Table S2 because samples are stratified to those positive (Ct < 28) by both qPCR and nl-qPCR for each target

**TABLE S5** Performance metrics for machine learning models

| Model | Ten-fold cross validation | Testing |
|---|---|---|
| Random Forest | | |
| ROC-AUC[a] | 0.637 ± 0.074 | 0.702 |
| Accuracy[a] | 0.586 ± 0.053 | 0.631 |
| PPV[a] | 0.674 ± 0.050 | 0.563 |
| NPV[a] | 0.497 ± 0.056 | 0.702 |
| F1 score[a] | 0.648 ± 0.048 | 0.632 |
| MCC[a] | 0.172 ± 0.104 | 0.263 |
| XGBoost | | |
| ROC-AUC[a] | 0.658 ± 0.080 | 0.687 |
| Accuracy[a] | 0.621 ± 0.043 | 0.627 |
| PPV[a] | 0.715 ± 0.048 | 0.567 |
| NPV[a] | 0.526 ± 0.048 | 0.686 |
| F1 score[a] | 0.647 ± 0.069 | 0.618 |
| MCC[a] | 0.241 ± 0.087 | 0.253 |
| Catboost | | |
| ROC-AUC[a] | 0.643 ± 0.084 | 0.686 |
| Accuracy[a] | 0.585 ± 0.062 | 0.632 |
| PPV[a] | 0.681 ± 0.064 | 0.582 |
| NPV[a] | 0.488 ± 0.064 | 0.679 |
| F1 score[a] | 0.619 ± 0.071 | 0.610 |
| MCC[a] | 0.169 ± 0.123 | 0.262 |
| K-Nearest Neighbors | | |
| ROC-AUC[a] | 0.607 ± 0.085 | 0.652 |
| Accuracy[a] | 0.561 ± 0.054 | 0.554 |
| PPV[a] | 0.635 ± 0.038 | 0.488 |
| NPV[a] | 0.539 ± 0.113 | 0.645 |
| F1 score[a] | 0.713 ± 0.047 | 0.600 |
| MCC[a] | 0.145 ± 0.125 | 0.120 |
| Logistic Regression | | |
| ROC-AUC[a] | 0.620 ± 0.056 | 0.681 |
| Accuracy[a] | 0.564 ± 0.055 | 0.625 |
| PPV[a] | 0.659 ± 0.054 | 0.569 |
| NPV[a] | 0.465 ± 0.054 | 0.680 |
| F1 score[a] | 0.603 ± 0.051 | 0.611 |
| MCC[a] | 0.126 ± 0.108 | 0.249 |

[a] ROC-AUC is the area under the curve of the receiver operating curve. Balanced accuracy was calculated as the average sensitivity and specificity of the model. PPV is the positive predictive value (PPV) and the metric for precision. NPV is the negative predictive value. The F1 score is calculated as the harmonic mean of precision (PPV) and recall. MCC is Matthews Correlation Coefficient..

**TABLE S6** Summary of input features for machine learning models

| Feature | Unit | Levels or ranges |
|---|---|---|
| Clinical | | |
| Self-reported antibiotic exposure | - | Yes or no |
| Dehydration status | - | No dehydration, moderate dehydration, severe dehydration |
| Emesis | - | Yes or no |
| Duration of diarrhea | Hours | [0, 312] |
| Sociodemographic | | |
| Age | Months | [1, 1080] |
| Sex | - | Male or female |
| Sample site | - | Habiganj, Naogaon, Patuakhali, Chattogram, Cumilla, or Kusthia |
| Microbiological | | |
| ICP1 detection (PCR)[a] | - | Positive or negative |
| ICP3 detection (PCR)[a] | - | Positive or negative |
| ICP1 quantity (qPCR Ct) | Ct values | [9.0, 35.3] |
| ICP1 detection (qPCR)[a] | - | Positive or negative |
| ICP3 quantity (qPCR Ct) | Ct values | [16.2, 35.9] |
| ICP3 detection (qPCR)[a] | - | Positive or negative |
| ICP1 quantity (nl-qPCR Ct)[a] | Ct values | [10.0, 35.0] |
| ICP1 detection (nl-qPCR)[a] | - | Positive or negative |
| ICP3 quantity (nl-qPCR Ct)[a] | Ct values | [17.7, 35.0] |
| ICP3 detection (nl-qPCR)[a] | - | Positive or negative |

[a] Features were only included in the feature ranking random forest model and were excluded from downstream models due to their low ranking.

**Table S7** Hyperparameter tuning of machine learning models

| Model | Hyperparameter | Values |
|---|---|---|
| Random Forest | 'n_estimators' | 10-100 |
| | 'max_depth' | 3-20 |
| | 'min_samples_split' | 2-10 |
| | 'min_samples_leaf' | 1-10 |
| | 'criterion' | 'gini' or 'entropy' |
| | 'bootstrap' | 'True' or 'False' |
| XGBoost | 'n_estimators' | 100-500 |
| | 'max_depth' | 3-15 |
| | 'learning_rate' | 0.001-1.0 |
| | 'gamma' | 0.0-1.0 |
| | 'subsample' | 0.6-1.0 |
| | 'colsample_bytree' | 0.6-1.0 |
| | 'objective' | 'binary:logistic' |
| | 'min_child_weight' | 1-10 |
| Catboost | 'iterations' | 50-500 |
| | 'depth' | 3-10 |
| | 'learning_rate' | 0.001-0.1 |
| | 'l2_leaf_reg' | 1.0-10.0 |
| K nearest neighbors (kNN) | 'n_neighbors' | 1-50 |
| | 'weights' | 'uniform', or 'distance' |
| Logistic regression (LR) | 'C' | 0.0001-10000 |
| | 'solver' | 'liblinear', 'saga', or 'lbfgs'[a] |
| | 'penalty' | 'l1' or 'l2'[a] |

[a] The 'liblinear' and 'saga' solvers were tested with both 'l1' and 'l2' options for penalty. However, the 'lbfgs' solver can only support the 'l2' penalty.
